# Evaluation and risk communication of effects of alcohol exposure on disposable procedure masks and portable air purifiers

**DOI:** 10.1101/2022.04.07.22273564

**Authors:** Yo Ishigaki, Shinji Yokogawa, Tatsuo Kato

**Author notes:** Corresponding author: Yo Ishigaki.

## Abstract

As electret technology can drastically improve the filtration efficiency of disposable procedure masks and portable air purifiers, it is widely used to prevent the airborne transmission of SARS-CoV-2. Furthermore, alcohol disinfectants are now widely used in offices, hospitals, and homes to prevent contact infection; hence, there is a concern that alcohol exposure may inactivate electret. In this study, 5 types of high-efficiency particulate air filter (HEPA) air purifiers—of which, one was made of fiberglass HEPA filter—14 types of cubical masks, and 11 types of pleated masks available to Japanese citizens were subjected to discharge according to the alcohol exposure protocol based on ISO (International Organization for Standardization) 16890, and changes in filtration efficiency and pressure drop were measured before and after the discharge. The results revealed that 17 (68%) of the 25 masks exhibited a significant decrease in filtration efficiency; this decrease due to discharge depended on the filter material. However, masks of polypropylene, polyethylene, and poly-vinylidene-difluoride composite fiber materials exhibited no significant decrease in filtration efficiency. In addition, 4 (80%) of the 5 HEPA filters showed a 40–64% decrease in filtration efficiency, while no decrease in filtration efficiency was observed for the fiberglass HEPA filter. Our survey (n = 500 Japanese adults, including 30 healthcare professionals) revealed that approximately 90% of the general public was unaware that the performance of masks and air purifiers could be degraded by direct spraying of alcohol—for disinfection purposes—or vapor exposure. Furthermore, 36% of the surveyed healthcare professionals indicated that they had sprayed alcohol directly on their masks. Therefore, based on the results of this experiment, we examined effective consumer warnings that could be utilized on the product labels and in the instructions. The results showed that it would be more effective to detail the extent and duration of the adverse effects of disregarding the precautions.

## 1. Introduction

Owing to the global spread of COVID-19, electret filters made of polymeric fibers such as polypropylene are widely used to prevent droplet or airborne transmission. It has been revealed—from the simulations using the supercomputer Fugaku—that non-woven masks are more efficient in collecting droplets than urethane or cloth masks (The Mainichi, 2021); hence, the Japanese Cabinet Secretariat has widely encouraged the public to wear non-woven masks to prevent COVID-19 infection (Prime Minister’s Office of Japan, 2021). A typical surgical mask consists of three layers, with the middle layer being a melt-blown non-woven fabric made of electretized polypropylene fibers, and the back and front protective layers are often spunbonded non-woven fabrics. N95 masks may also combine electretized needle-punched non-woven fabrics (Ishigaki, 2022).

Ou et al. (2020) used actual medical surgical masks sold by 3M and Halyard and N95 masks to determine the effectiveness of the N95 masks. The authors reported that immersion into or direct spraying of IPA (isopropyl alcohol) eliminated electrostatic forces and significantly reduced filtration efficiency to unacceptable levels. The US Centers for Disease Control and Prevention (CDC) offer the following decontamination methods rather than alcohol for N95 masks: chlorine dioxide, sodium chlorite, supercritical or dense-phase CO_2_, dry heat sterilization, steam sterilization, ultraviolet radiation, ozone, peracetic acid, plasma sterilization, hydrogen peroxide, electron beam irradiation, laundry drying, and methylene blue (CDC, 2021).

Meanwhile, Japan’s Ministry of Health, Labor and Welfare recommends the installation of portable air purifiers equipped with HEPA filters to reduce the risk of airborne infection in poorly ventilated spaces, based on the recommendations of the American Society of Heating, Refrigerating and Air-Conditioning Engineers (ASHRAE, 2020), the Federation of European Heating, Ventilation and Air Conditioning associations (REHVA, 2020), and the CDC (2003). Nishimura (2011) found that portable air purifiers from four major Japanese manufacturers: Panasonic, Daikin, Sharp, and Mitsubishi, were used in a 14.4-m^3^ chamber sprayed with influenza virus cultures. Qian et al. (2010) reported that the HEPA air purifiers reduced the number of airborne viruses below the detection limit, whereas the non-HEPA air purifiers only reduced the number of viruses faster than natural decay. The portable HEPA air purifier was tested in operation in a larger 108.5 m^3^ laboratory and yielded an air purification capacity of 2.5–5.6 air purification cycles per hour in terms of ventilation frequency. Against this backdrop, the use of HEPA is widely recommended in Japan for the prevention of airborne diseases.

In Japan, HEPA is defined by the Japan Industrial Standard (JIS) Z8122 as “an air filter with a particle filtration efficiency of 99.97% or higher for particles of 0.3 µm in diameter at rated airflow and an initial pressure drop of 245 Pa or less. However, the IPA treatment discharging process specified in ISO 16890 is not applied in the performance evaluation of HEPA filters specified in JIS Z8122. Therefore, many portable air purifier manufacturers use non-woven electret filters as HEPA filters. Generally, non-woven HEPA has a lower pressure drop than glass fiber HEPA, making it easier to achieve a large airflow in a compact housing area, hence, rendering it suitable for use in the compact Japanese houses. However, like masks, electret is deactivated upon exposure to alcohol (Schuldt et. al. 2020).

As mentioned above, electret filters are widely used in masks and portable air purifiers. However, their vulnerability to alcohol is not well known to the public. In a web-based survey of 500 Japanese adults (246 males and 254 females, mean age of 49 years), 50 (10%) and 45 (9%) of them were aware that alcohol atomization degrades the filter performance of masks and air purifiers, respectively. In addition, 105 (21%) and 25 (5%) of the respondents had sprayed rubbing alcohol directly on their masks of and air purifiers, respectively; in such cases where the general public unintentionally deactivates electret, they are at risk of using the product under lower filtration performance.

Furthermore, evaluation of the response trends of healthcare professionals, who comprised 30 (6%) of the 500 respondents, revealed that the number of individuals who had sprayed alcohol on their masks was significantly higher among the healthcare professionals (p = 0.0377* in Fisher’s exact test for contingency tables) at 36%, and 20% for non–healthcare professionals. This may be because people were forced to disinfect their masks for reuse when the COVID-19 epidemic caused a worldwide shortage of masks.

The probability of having sprayed alcohol on portable air purifiers was also significantly higher among healthcare professionals (p = 0.0105* in Fisher’s exact test), at 16% for healthcare professionals and 4% for non–healthcare professionals. This suggests that some medical facilities are forced to frequently disinfect areas—where hands come in contact—with alcohol.

Conversely, 23% of healthcare professionals, as opposed to 9% of non-healthcare professionals, understood that spraying alcohol on masks irreversibly reduces the masks’ performance (p = 0.0219* in Fisher’s exact test); similarly, 26% of healthcare professionals, as opposed to 8% of non-healthcare professionals, were aware that the performance of portable air purifiers is irreversibly impaired upon exposure to alcohol (p = 0.0025* for Fisher’s exact test). Furthermore, less than 25% of the healthcare professionals knew that alcohol inactivates electret, indicating the need for increased awareness.

Sugihara (2021) and Band et. al. (2021) proposed the use of a Van de Graaff electromotive force generator to re-charge masks for reuse, and a microwave oven to re-activate electrets that have been deactivated by alcohol exposure. However, performing such a process in ordinary homes and hospitals would be impractical. Therefore, the aforementioned risk should be appropriately communicated in product packaging and guidelines.

In this study, we measure the degree of reduction in filtration efficiency using 11 types of pleated masks, 14 types of cubical masks, and 5 types of HEPA air purifiers—available to the Japanese public—through an alcohol exposure test based on ISO 16890. Based on the results, effective communication stating and highlighting the risk to consumers is discussed.

## 2. Material and methods

The filter samples comprised 14 cubical mask products shown in Table 1, 11 pleated mask products shown in Table 2, and filters for 5 HEPA-type portable air purifiers shown in Table 3. Each sample was discharged by exposure to a saturated atmosphere of IPA for 24 h followed by exposure to air for 1 h. Samples were evaluated for filtration efficiency before and after discharging using the measurement system shown in Figure 1. The measurement system comprised an exhaust blower (VFC308PN, Fuji Electric, Kanagawa, Japan), a flow meter (25-250 l/min, Nippon Flow Cell, Tokyo, Japan), a digital manometer for measuring the differential pressure between the primary and secondary sides (QDP33, Yamamoto Electric Works, Hyogo, Japan), and two particle counters (KC-22B and KC-01E, Rion, Tokyo, Japan). The flow rate during the test was one pattern of 28.3 l/min for the masks and four patterns of 36, 72, 117, and 234 l/min for the HEPA filters.

**Table 1.** specification for cubical masks (PP: polypropylene, PEs: polyester, PE: polyethylene, PVDF: polyvinylidene difluoride, PLA: poly-lactic acid, PU: polyurethane, PET:polyethylene terephthalate)

| ID | Manufacturer / Seller | Brand / Product name / model | Main filter material(s) |
| --- | --- | --- | --- |
| C1 | AIRINUM | LITE AIR FILTER-OPTIMAL<br>(LITE AIR MASK) | PP, PEs |
| C2 | AIRINUM | URBAN AIR FILTER 2.0<br>(URBAN AIR MASK 2.0) | PP, PEs |
| C3 | PHILIPS | PHILIPS Breeze Mask<br>ACM066 | PP, PEs |
| C4 | Jiangxi AICARE Medical<br>Technology Co., Ltd. | PROTECTIVE MASK KN95 | PP |
| C5 | Yagi & Co., Ltd. | AirQueen | PP, PE,<br>PVDF |
| C6 | TBM Co. | Bio Face | PLA |
| C7 | Samurai Works, Inc. | Nano Cool Antibacterial Mask | PP, PEs |
| C8 | Mitsufuji Corp. | 100 times washable summer<br>mask hamon AG | PEs |
| C9 | Molecular Labs, Inc. | Molecular mask | Cotton,<br>PET |
| C10 | Durio PPE Sdn Bhd | Durio KN95 | PP |
| C11 | KYUNGIN FLAX Co., Ltd. | FLAX KF94 | PP |
| C12 | Avanti Corporation | LALA KF94 High performance<br>masks | PP |
| C13 | Pioneer System Co. | Pioneer Mask | PP |
| C14 | ARAX Co., Ltd. | PITTA MASK | PU |

**Table 2.** specifications for pleated masks

| ID | Manufacturer / Seller | Brand / Product name / model | Main filter material(s) |
| --- | --- | --- | --- |
| P1 | First Rate | FR-185 | PP |
| P2 | AEON | TOPVALU Non-woven fabric<br>mask for virus droplets and<br>PM<sub>2.5</sub> | PP, PE |
| P3 | IRIS OHYAMA | Surgical mask | PP |
| P4 | Sansyo | Sun clean mask SCM-1 | PP |
| P5 | System Polymer Co., Ltd. | System Polymer Non-Woven Omega Type Pleated | PP |
| P6 | Ace International Corp. | MORSE GUARD | PP |
| P7 | Dandong Tianhao Clarification Material Co., Ltd. | Single-use masks (Green) | PP |
| P8 | BYD Co., Ltd. | BYD Electronics | PP |
| P9 | Craftsman Co. | 2-layer nonwoven fabric mask S-011 | PP |
| P10 | System Polymer Co., Ltd. | System Polymer 3-Layer Non-Woven Omega Type Pleated | PP |
| P11 | Delta Electronics (Japan), Inc | Delta Mask | PEs, PE, PP |

**Table 3.** Specifications of HEPA filters in portable air purifiers

| ID | Manufacturer / Seller | Brand / Product name / model | Main filter material(s) |
| --- | --- | --- | --- |
| H1 | SHINWA | VG-HEPA | Glass fiber HEPA |
| H2 | SHARP | FZ-E75HF | Electret HEPA |
| H3 | DAIKIN | KAFP080B4 | Electret HEPA |
| H4 | Panasonic | F-ZXLP90 | Electret HEPA |
| H5 | Blueair | F200300PA | Electret HEPA |

**Figure 1.**
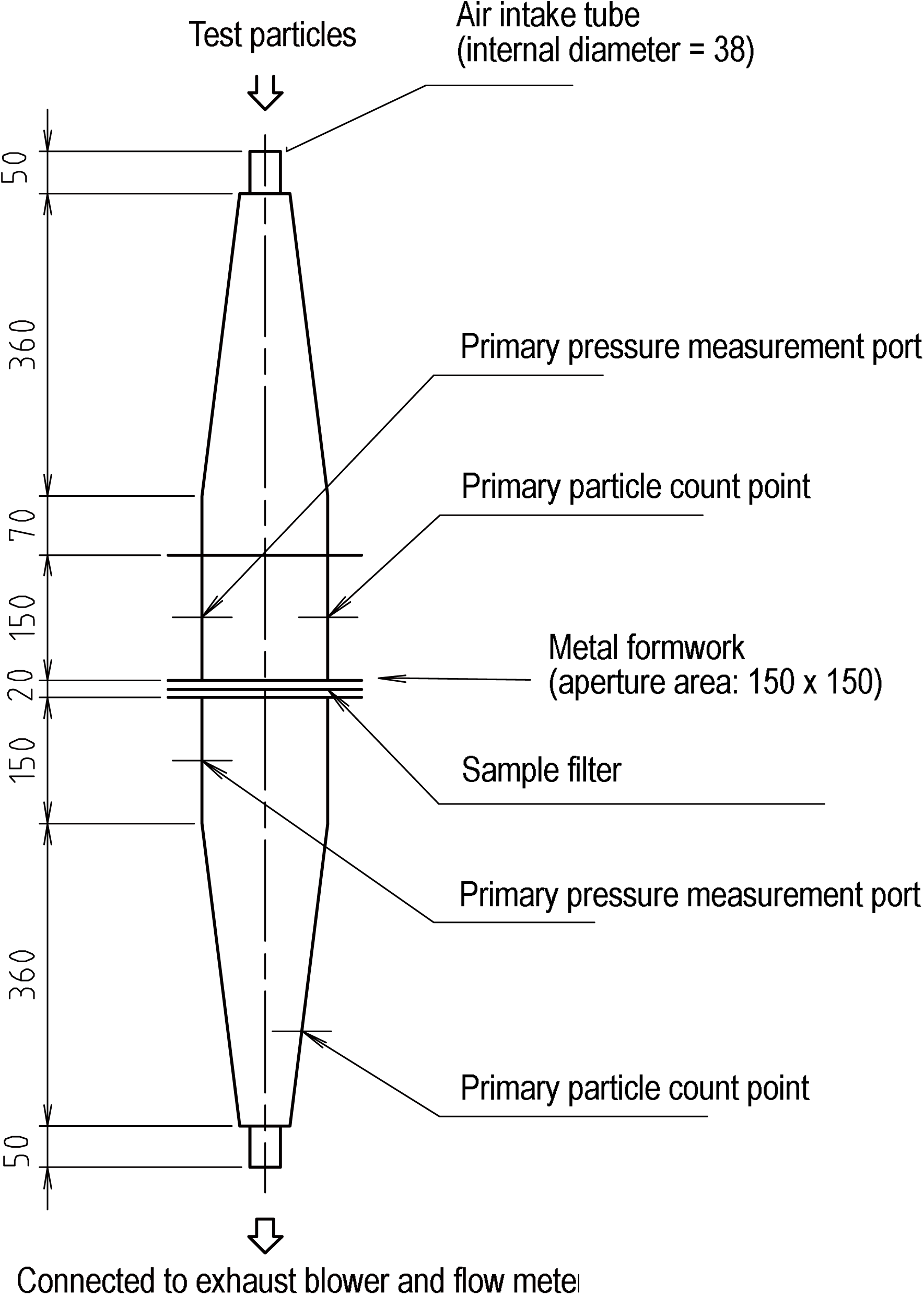
Measurement system for filtration efficiency and differential pressure (side view)

Test particles comprised background particles naturally occurring in the atmosphere in the case of the mask, and test particles with center diameters of 0.122, 0.173, 0.240, 0.387, and 0.700 µm in the case of the HEPA filter. Both masks and HEPA filters were set in a solid formwork with caulking material to prevent air leakage. The formwork had a 150 × 150 mm square aperture; the cubical mask was set in a formwork unfolded on a flat surface, and the pleated mask was set in a formwork with its pleats unfolded and stretched. The HEPA filter was set in a formwork without deforming the filter’s bellow shape. As the seal plate was installed, the effective filtration area for HEPA was 10,000 mm^2^.

## 3. Results and considerations

Figure 2 reveals the measured pressure drop across the masks before and after discharge. Of the 25 samples, 2 samples exhibited an increase of >10% in pressure drop after the discharge, whereas 6 samples showed a decrease. The large decrease in pressure drop in P11 was attributed to the fact that the filter of this mask was a composite of polyester (PEs), Polyethylene (PE), and Polypropylene (PP), and that alcohol added some binders. This may have changed and affected the fiber structure.

**Figure 2.**
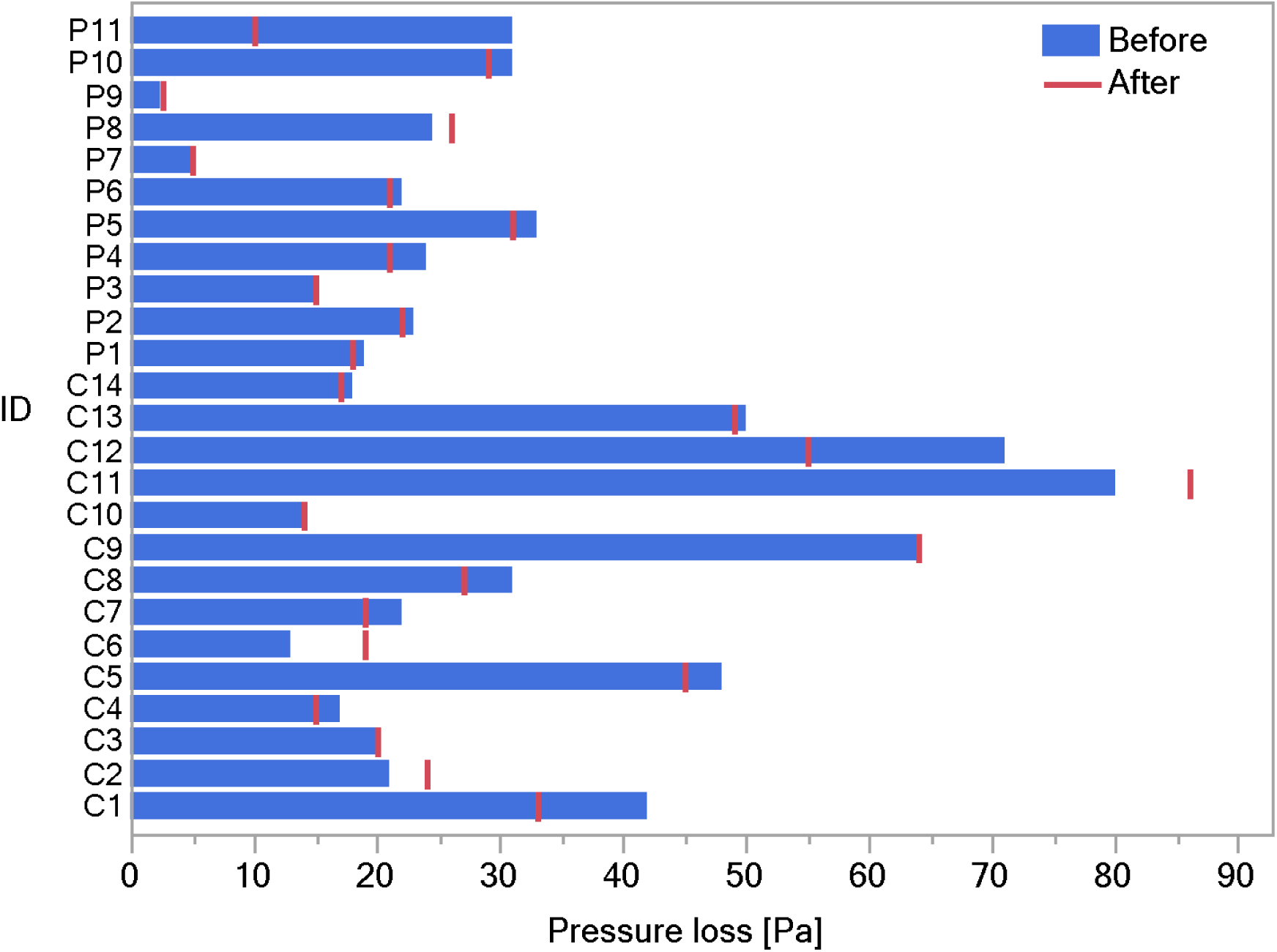
Pressure drop across the masks before and after discharge

One reason underlying the large increase in the pressure drop at C6 was that the main material of the filter in this mask was Poly-Lactic Acid (PLA); Gondo et al. (2011) have reported that this material exhibits alcohol solvent-induced crystallization upon exposure to PLA. As pointed out in the previous section, the discharge process may have caused compositional changes in the filter fibers.

Figure 3 reveals the measured filtration efficiency of the masks before and after discharge. Out of the 25 samples, 17 (C1–C4, C10–C13, P1–P7, and P10–P11; 68% of the total) exhibited a significant decrease in filtration efficiency due the discharge process. All these samples contained polypropylene in the filter material, and were therefore, assumed to be using an electret filter. In addition, filtration efficiency tended to decrease for C6, which was mainly made of PLA. In principle, PLA can be processed into an electret filter by external corona discharge after being formed as a melt-blown nonwoven fabric (Zhang et al., 220); therefore, C6 may also be an electret filter. The pressure drops in C6 and P11 also changed by more than 30%, suggesting that the compositional change in the filter fibers caused by the discharge process may have affected the filtration efficiency.

**Figure 3.**
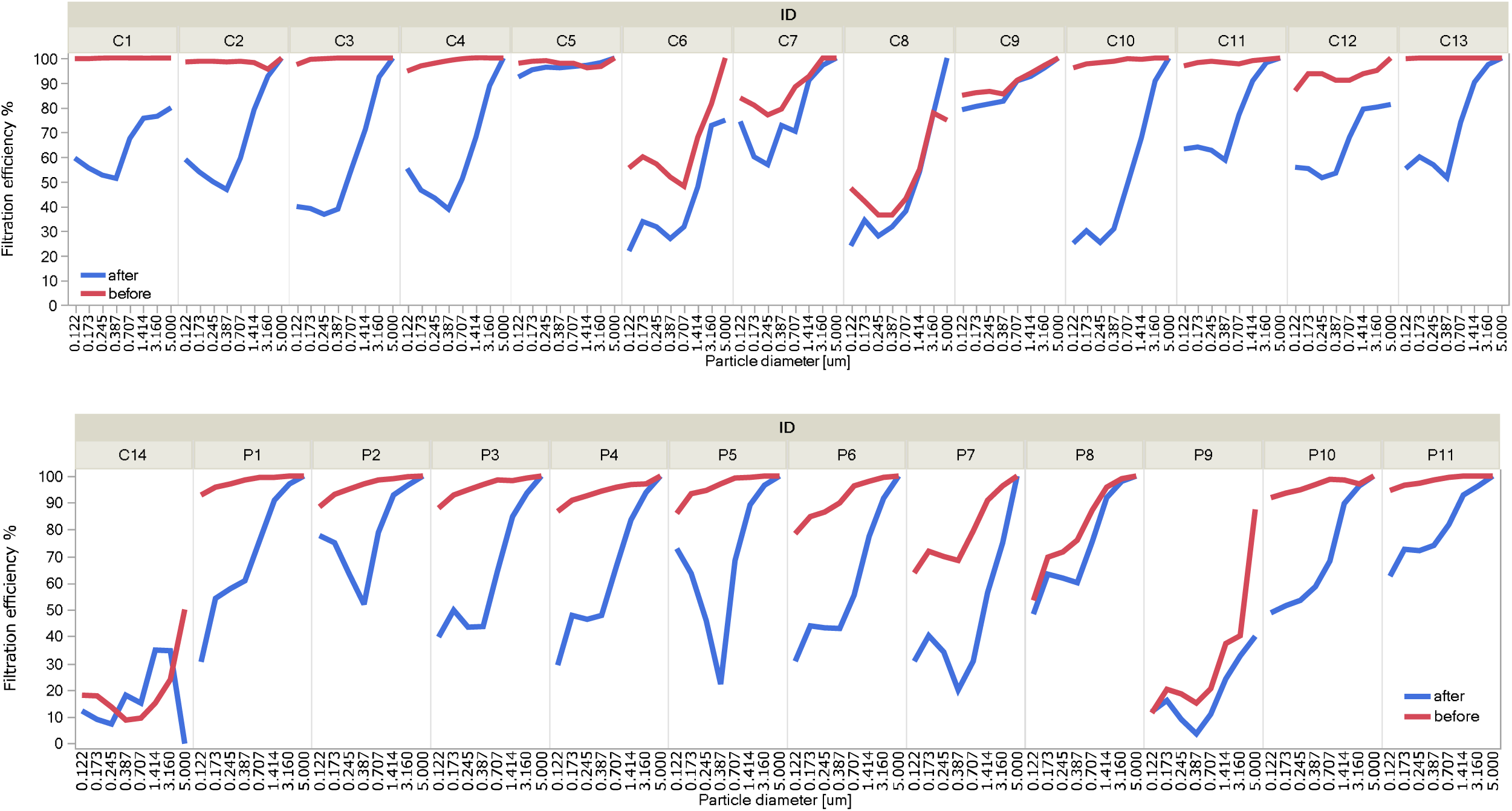
Filtration efficiency of the masks before and after discharge

Conversely, the effect of discharge was limited for the remaining C5–C9, C14, and P8–P9; because C5, C8–C9, and C14 were made of PVDF, PE, cotton, PU, or PET (and not polypropylene), which were considered non-electret filters. C14 exhibited minimal filtration efficiency because of its material, a porous sponge made of polyurethane. Although P8 and P9 were made of polypropylene, they may be non-electret filters or less active as electret filters owing to their low original filtration performance before discharge. As P9 had a two-layer structure, the middle layer of melt-blown non-woven fabric, which is usually used as an electret filter, was possibly omitted.

The material of each sample of mask filters, the pressure drops due to discharge, and the change in filtration efficiency for each particle size were interdependent. It was also necessary to consider the effect of inter-individual differences in the samples used. Therefore, using the generalized linear mixed model, we fit a mixed model with particle size, with and without discharge, shape (C or P), material type, and the interaction between material type and discharge as explanatory variables to obtain fixed effects. Logit transformation was used for filtration efficiency, and log transformation was used for particle size. The random effect, which indicates inter-individual differences, was p = 0.0097, indicating that inter-individual differences and fixed effects could be separated.

Most of the fixed effects were considered significant (p < 0.05): particle size (logarithm), presence of discharge, certain material groups, and the interaction between material type and discharge (material discharge). The estimated results for each parameter (Table 4) reveal that the filtration efficiency increases as the particle size increases and that the filtration efficiency decreases with discharge. The *interaction between material and discharge* revealed a significant decrease in filtration efficiency due to discharge (p < 0.01) for PEs/PE/PP, PP, PP/PE, and a tendency to decrease for PP/PEs.

**Table 4.** Results of generalized linear mixed model analysis of filtration efficiency

| Term | Estimates | Standard Error | Degrees of freedom, denominator | t-value | p-value, prob>\|t\| |
| --- | --- | --- | --- | --- | --- |
| Intercept | 1.748750 | 0.314602 | 15.4 | 5.56 | **<0.0001\*** |
| logarithm of particle diameter | 0.635253 | 0.043786 | 299.6 | 14.51 | **<0.0001\*** |
| Discharge[after] | -0.713639 | 0.070898 | 299.4 | -10.07 | **<0.0001\*** |
| Shape[C] | 0.461801 | 0.272037 | 15.3 | 1.7 | 0.1099 |
| Material[Cotton/PET] | 0.360869 | 0.919562 | 15.1 | 0.39 | 0.7002 |
| Material[PEs] | -1.98356 | 0.918146 | 15 | -2.16 | **0.0474\*** |
| Material[PEs/PE/PP] | 2.049794 | 0.988099 | 15.4 | 2.07 | 0.0552 |
| Material[PLA] | -1.77459 | 0.918146 | 15 | -1.93 | 0.0724 |
| Material[PP] | 0.592671 | 0.417984 | 15.2 | 1.42 | 0.1763 |
| Material[PP/PE] | 1.708347 | 0.983659 | 15.1 | 1.74 | 0.1028 |
| Material[PP/PE/PVDF] | 1.849194 | 0.919562 | 15.1 | 2.01 | 0.0626 |
| Material[PP/PEs] | 0.623883 | 0.545927 | 15.4 | 1.14 | 0.2705 |
| Material[Cotton/PET]*Discharge[after] | 0.571799 | 0.218259 | 299.2 | 2.62 | **0.0092\*** |
| Material[PEs]*Discharge[after] | 0.574745 | 0.212238 | 299.3 | 2.71 | **0.0072\*** |
| Material[PEs/PE/PP]*Discharge[after] | -0.62455 | 0.237200 | 299.2 | -2.63 | **0.0089\*** |
| Material[PLA]*Discharge[after] | 0.240765 | 0.212095 | 299.2 | 1.14 | 0.2572 |
| Material[PP]*Discharge[after] | -0.56157 | 0.090994 | 299.7 | -6.17 | **<0.0001\*** |
| Material[PP/PE]*Discharge[after] | -0.36977 | 0.218259 | 299.2 | -1.69 | 0.0913 |
| Material[PP/PE/PVDF]*Discharge[after] | 0.400809 | 0.218259 | 299.2 | 1.84 | 0.0673 |
| Material[PP/PEs]*Discharge[after] | -0.99995 | 0.139130 | 301.2 | -7.19 | **<0.0001\*** |

Figure 4 plots the filtration efficiency before and after discharge for each material type; the filtration efficiency before discharge is relatively high for PEs/PE/PP, PP, PP/PE, PP/PE/PVDF, and PP/PEs, and relatively low for PEs, PLA, and PU. The only material with relatively high filtration efficiency and no significant decrease in filtration efficiency due to discharge was PP/PE/PVDF used in C5.

**Figure 4.**
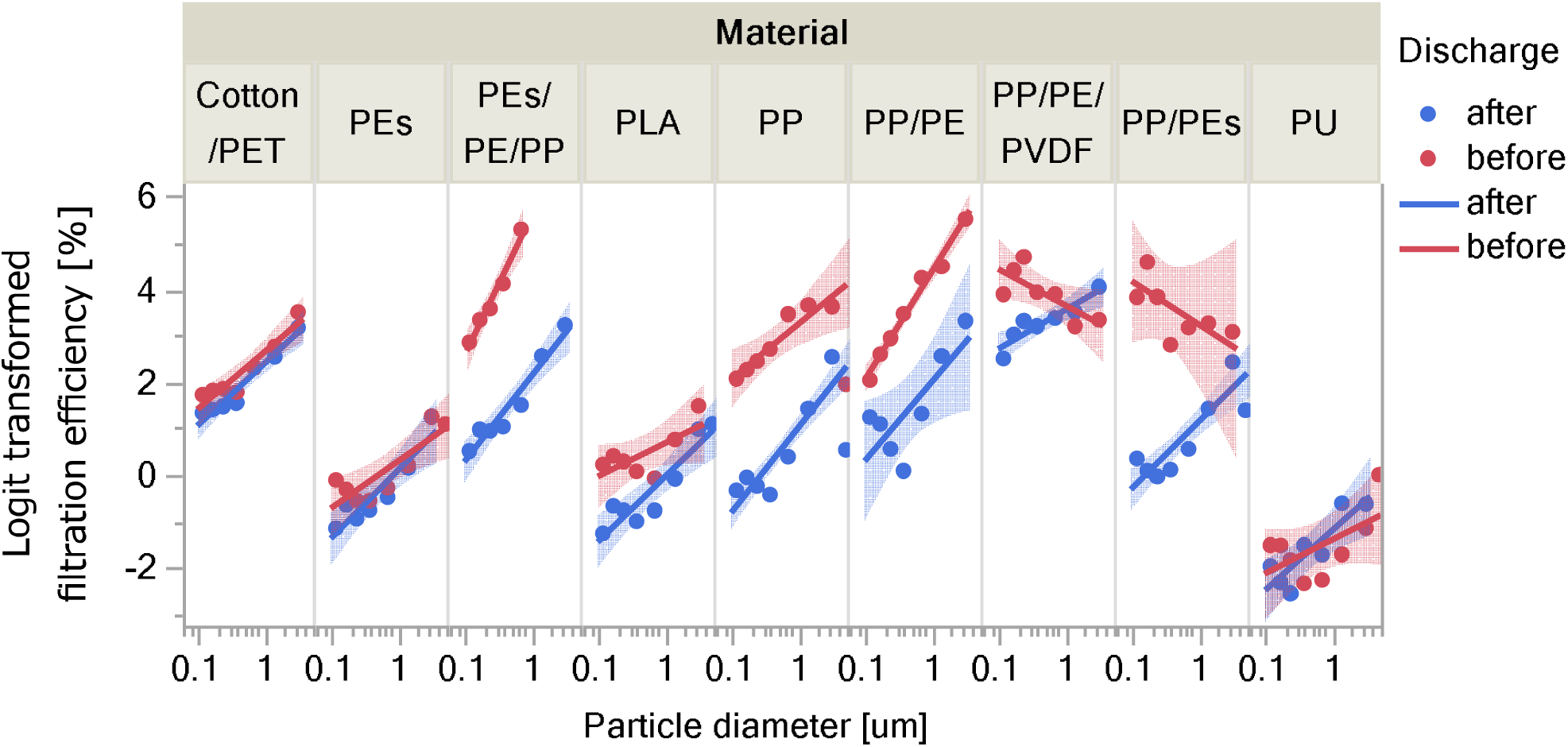
Filtration characteristics before and after discharge for each material (The filled areas indicate 95% confidence intervals. Particle size was logit-transformed; thus 100% collection efficiency was not plotted)

Figure 5 reveals the measured pressure drop across the HEPA filter before and after discharge. In both samples, the amount of change in pressure drop at each airflow was less than 5 percent point. In the mask experiment, two samples were found to exhibit large changes in pressure drop that could be attributed to compositional changes, but no such changes were found in the four HEPA filters used in this experiment.

**Figure 5.**
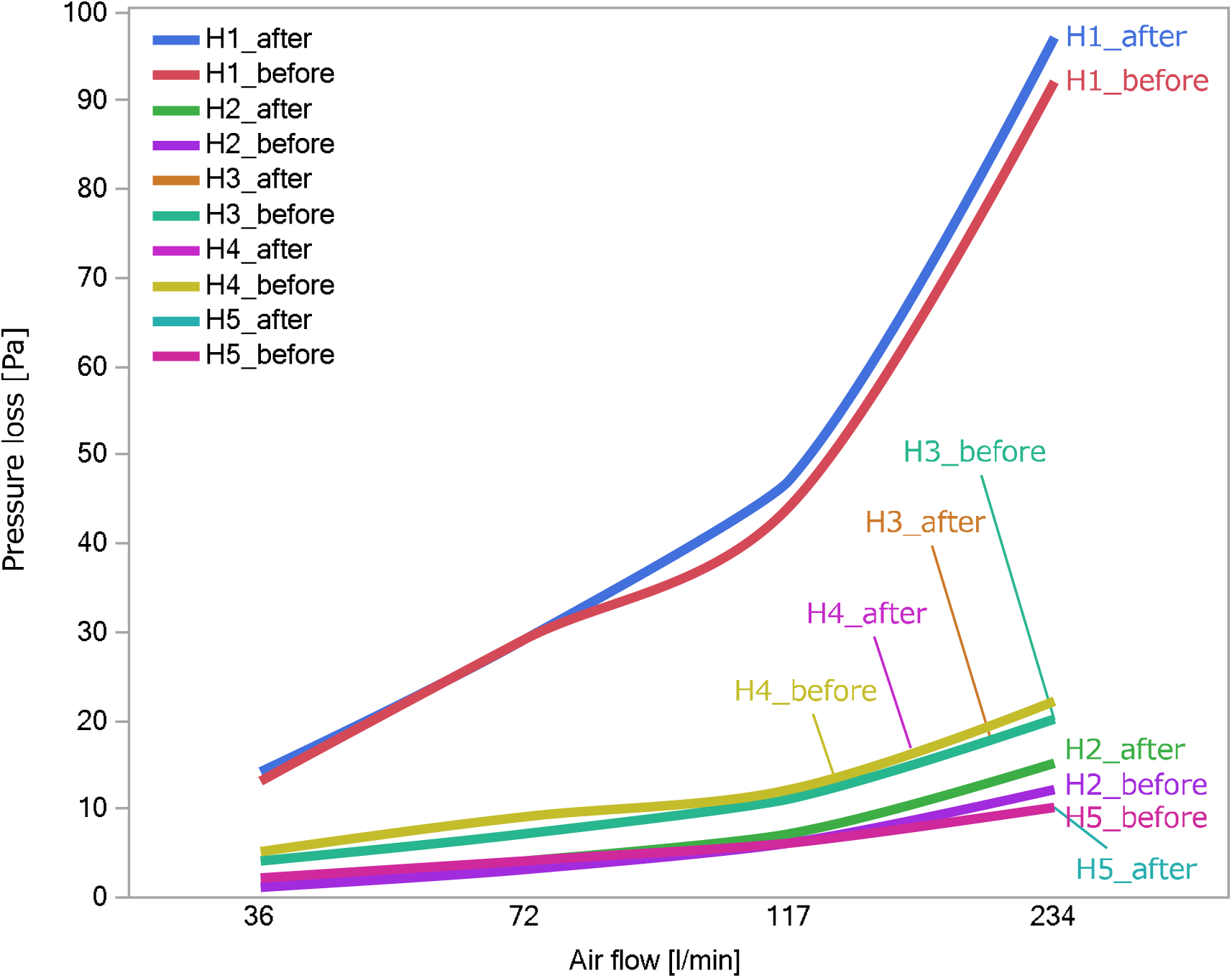
Pressure drop across the HEPA filters before and after discharge (complemented by spline curve)

Figure 6 reveals the measured filtration efficiency of the HEPA filters before and after discharge; all samples had almost 100% filtration efficiency before discharge, given that they are HEPA filters. The average reduction was 0, 40, 53, and 64% for H1, H2, H3, and H4, respectively. Among these filters, only H1 was made of fiberglass, while the other samples were all electret filters, suggesting that only H1 was unaffected by the discharge.

**Figure 6.**
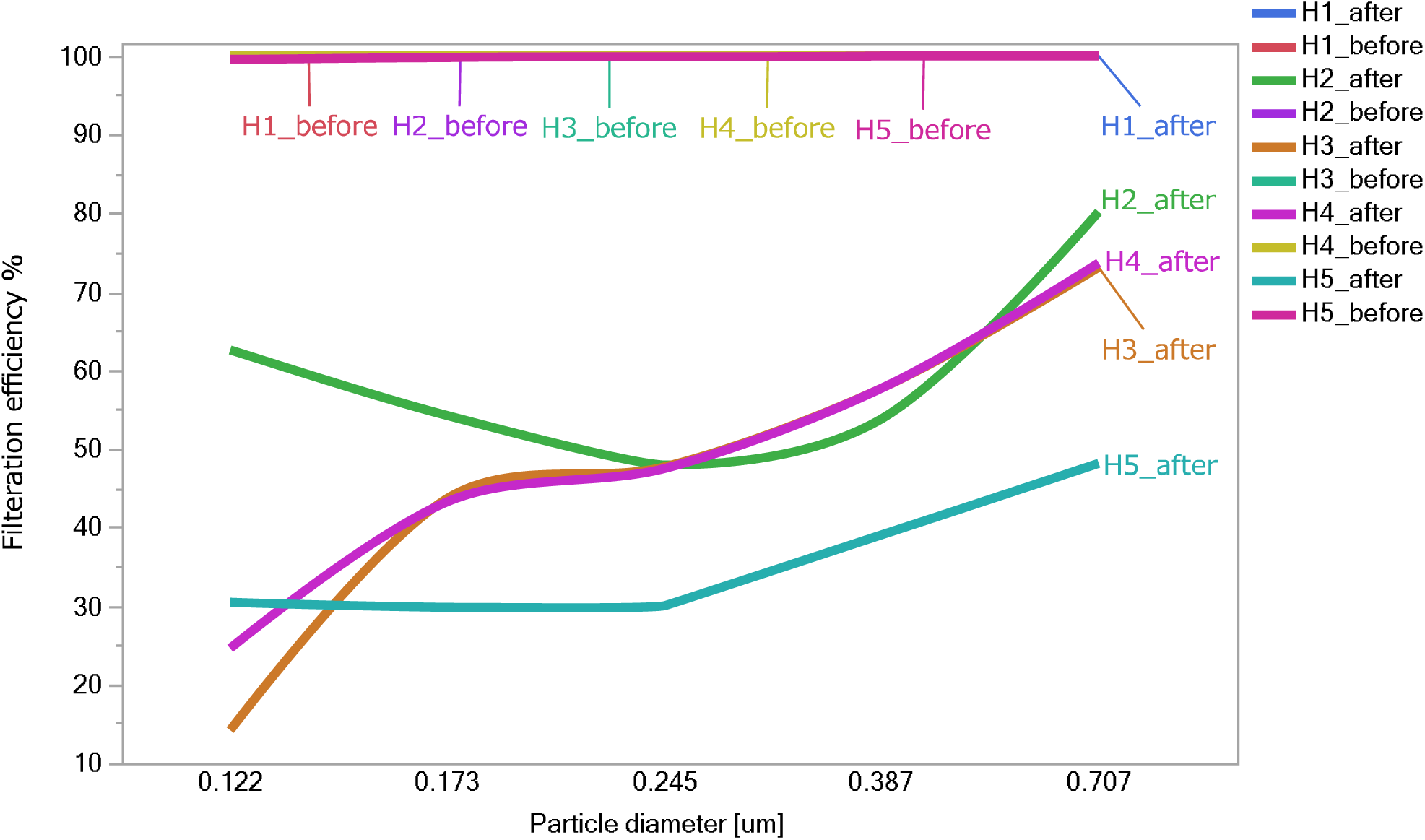
Filtration efficiency of the HEPA filters before and after discharging (Complemented by spline curve)

Even after discharge, H2 maintained a filtration efficiency of >50% compared to H3, H4, and H5, especially for small particle sizes below 0.173 µm.

## 4. Discussion

### 4-1 Regarding mask efficiency

Generally, except for electrostatic collection, the collection principles of filters for particles of micron order or smaller can be classified into three categories: diffusion, interception, and impaction. Of these, diffusion contributes to relatively small particles, while interception and impaction contribute to relatively large particles (Lindsley, 2016). Therefore, H2 may have used a polypropylene material with a finer fiber diameter or had a higher fill rate than the other filters, which may have contributed to Brownian diffusion.

As H3 and H4 exhibited similar filtration efficiency characteristics after discharge, it can be inferred that the fiber diameter and filling ratio were also similar.

The average decrease in filtration efficiency after discharge was the highest for H5, which is thought to be due to the ionizer originally installed in front of the filter, which charges the passing particles and increases the collection effect by electrostatic charge. This means that compared to other filters, a higher percentage of the filters rely on the electrostatic collection mechanism, and the mechanical filtration efficiency of the filter may be low.

### 4-2 Regarding risk of communication

The discharge process resulted in a decrease in filtration efficiency in 68% of the mask samples, which was inferred to be typical of electret filters. Similar post-discharge results were observed for all electret HEPA filters, with an average of 40–64% reduction in filtration efficiency. These results indicate that if the public unintentionally deactivates the electret using alcohol, users are at risk of unknowingly using less efficient products.

Therefore, we provided five types of risk communication messages shown in Table 5 and asked the 500 Japanese adult subjects to select the message that was more effective for changing behavior if it was displayed as a warning on the product package or in the instruction manual. The results are presented in Table 5.

**Table 5.** Examples of warning signs and the percentage of users who find them effective

| No. | Message | % |
| --- | --- | --- |
| 1 | Do not use rubbing alcohol. | 15.4 |
| 2 | Do not use rubbing alcohol near the product. | 10.4 |
| 3 | Do not use rubbing alcohol near the product as it will degrade performance. | 21.6 |
| 4 | Do not use rubbing alcohol near the product as it will degrade performance and prevent restoration. | 23 |
| 5 | Do not use rubbing alcohol near the product as it will irreversibly decrease the filtration efficiency by 50%. | 29.6 |

The risk communication messages are designed to progressively be more specific: No. 1 simply states the prohibition; No. 2 adds information on the location of the alcohol use; No. 3 gives the reason for the prohibition; No. 4 explains the degree of temporal impact in relatively greater detail; and No. 5 quantifies the degree of performance deterioration. The results of the questionnaire survey indicate that the most informative message, No. 5, is appropriate as a warning.

The masks and portable air purifiers used in this experiment did not indicate such a warning message. However, because rubbing alcohol is used routinely, reminding people not to expose their products to alcohol is an important risk communication in infection control. Conversely, pictograms of such precautions would communicate the risk more extensively and inform multilingual users and those who are unable to read about these risks.

The questionnaire survey also revealed that healthcare professionals were significantly more likely to be aware of the performance deterioration due to alcohol exposure to electret filters. However, the spraying of alcohol on masks and portable air purifiers was performed by a higher number of healthcare professionals than non-healthcare professionals, probably because alcohol disinfection is required in all situations in the medical field. Therefore, if electret masks are used in work environments where frequent exposure to alcohol is inevitable, encouraging frequent mask changes may be beneficial. Alternatively, it may be effective to have workers wear masks made of materials that have relatively high filtration efficiency and are not subject to the effects of discharge, as observed in C5. When continuously operated portable air purifiers are used in medical facilities, fiberglass HEPA should be used instead of electret HEPA.

In addition to informing the general public, healthcare institutions should consider issuing new risk communication messages regarding electret filters. From an engineering standpoint, developing a practical device (Yoshida, 2021) that can simply check the activity of electret filters by measuring their thermally stimulated current would be beneficial.

In this experiment, the filter was exposed to an IPA-saturated atmosphere for only 1 day as per ISO 16890. However, to evaluate the effects in a real environment, additional testing by exposure of electret to lower concentrations of alcohol for relatively longer periods of time will be required in future research.

## 5. Conclusions

Alcohol-exposure tests of 25 disposable procedure masks available to Japanese citizens showed a significant decrease in filtration efficiency in 68% of the products. The masks that did not show a significant decrease were constructed from PP/PE/PVDF material. As noted by Ou et al. (2020), the filtration efficiency of medical and surgical masks decreases with alcohol exposure. The present study quantitatively demonstrates that the same risk applies to disposable procedure masks used daily by members of the general public.

Furthermore, alcohol exposure tests on five different HEPA filters in portable air purifiers showed that 80% of the products, whose filters are all electret HEPA type, had a 40–64% reduction in filtration efficiency, while glass fiber HEPA showed no reduction in filtration efficiency. The percentage reduction in efficiency for each particle size was characterized separately for each product, and possible influences of fiber diameter, fill factor, and ionizer were found.

Results of a web-based survey of 500 Japanese adults revealed that approximately 90% of respondents were unaware of the reduced-performance risk of air purifiers resulting from direct spraying of alcohol or vapor exposure, and 20% had sprayed alcohol on their masks. In addition, it was identified that healthcare professionals sprayed alcohol onto masks and portable air purifiers at a significantly higher rate. Accordingly, we examined the effectiveness of warning signs to inform consumers not to spray alcohol onto masks or portable air purifier products. We found it most effective to detail the extent and duration of adverse events that would occur if the precautions were not followed.

This study notably quantifies the vulnerability of masks and air purifiers to alcohol, identifies the lack of general knowledge about that vulnerability, and makes recommendations for effective risk warnings to consistently and effectively convey that knowledge. A weakness of this study is that the alcohol exposure test follows ISO 16890 rather than replicating the lower concentration and longer exposure conditions of actual medical facilities and households. This weakness will be overcome by additional future investigations.

## Data Availability

All data produced in the present study are available upon reasonable request to the authors.

## ETHICS AND REPORTING

## Acknowledgement

This study was approved (approval number 21079) by the Ethics Committee on Experiments on Human Subjects of the University of Electro-Communications, Chofu, Tokyo, Japan. The authors declare no conflicts of interest associated with this manuscript. This work was supported by a KAKENHI Grant (No. 21K19820) from JSPS.

